# Psychotic-like experiences and polygenic liability in the ABCD Study®

**DOI:** 10.1101/2020.07.14.20153551

**Authors:** Nicole R. Karcher, Sarah E. Paul, Emma C. Johnson, Alexander S. Hatoum, David AA Baranger, Arpana Agrawal, Wesley K. Thompson, Deanna M. Barch, Ryan Bogdan

**Affiliations:** Department of Psychiatry, Washington University School of Medicine, St. Louis, MO; Department of Psychological and Brain Sciences, Washington University, St. Louis, MO; Department of Psychiatry, University of Pittsburgh, PA; Division of Biostatistics, Department of Family Medicine and Public Health, University of California, San Diego

## Abstract

**Background:** Psychotic-like experiences (PLEs) during childhood are harbingers for severe psychopathology, including psychotic disorders, and neurodevelopmental impairments in adolescence and adulthood.

**Methods:** Data from children of genomically-confirmed European ancestries (n=4,650; ages 9-10; 46.8% female) who completed the baseline Adolescent Brain Cognitive Development⍰ Study session were used to assess whether PLEs (i.e., both total and the presence of significantly distressing) are associated with polygenic scores (PGS) related to psychopathology (i.e., schizophrenia [SCZ], educational attainment [EDU], psychiatric cross-disorder risk [CROSS], PLEs). We also assessed whether variability in global and region indices of brain structure (i.e., volume, cortical thickness, surface area) as well as behaviors proximal to PGS (e.g., cognition for EDU) indirectly linked PGS to PLEs using mediational models.

**Findings:** EDU and CROSS PGS were associated with total and significantly distressing PLEs (all %ΔR^2^s=0.202-0.660%; ps<0.002). Significantly distressing PLEs were also associated with higher SCZ and PLEs PGS (both %ΔR^2^=0.120-0.171%; ps<0.02). Global brain volume metrics and cognition indirectly linked EDU PGS to PLEs (proportion mediated: 3.33-32.22%).

**Interpretation:** Total and distressing PLEs were associated with genomic risk indices associated with broad spectrum psychopathology risk (i.e., EDU and CROSS PGS). Significantly distressing PLEs were associated with genomic risk for psychosis (i.e., SCZ, PLEs). Global brain volume metrics and PGS-proximal behaviors represent promising putative intermediary phenotypes that may contribute to genomic risk for psychopathology. Broadly, polygenic scores derived from genome-wide association studies of adult samples can generalize to indices of psychopathology risk among children and aid the identification of putative neural and behavioral intermediaries of risk.

**Funding:** National Institute of Health

## Introduction

Psychotic-like experiences (PLEs) are nonclinical schizophrenia spectrum symptoms that include perceptual abnormalities and mild delusional thoughts. They commonly occur in children (∼10% of children and adolescents) and are considered a dimensional, transdiagnostic marker of significant psychopathology risk (e.g., odds ratio ∼ three), including conversion to adult psychotic disorders in some children.^1,2^ The adverse mental health prognosis of children with PLEs, even beyond those who eventually develop schizophrenia, has inspired efforts to identify children at risk for their development and improve our understanding of PLE etiology to ultimately facilitate advances in prevention and treatment.

Building upon twin work documenting the moderate heritability of PLEs, genome-wide association studies (GWASs) have shown that PLEs are highly polygenic, much like other complex behavioral and biological phenotypes.^e.g.3^ Results from well-powered “discovery” GWASs may be projected onto individuals in an independent sample by averaging common variants - weighted by GWAS effect size and number of risk alleles present - across the genome to generate polygenic scores (PGS) that represent an individual’s genomic predisposition for the “discovery” GWAS phenotype.^4^ In contrast to initial evidence reporting null associations between schizophrenia PGSs and adolescent PLEs using a relatively low-powered “discovery” GWAS (cases n=9,394^5^),^e.g.,6^ recent work leveraging results from a large GWAS of schizophrenia (cases n=36,989^7^) has generally linked schizophrenia PGSs to PLEs during adolescence and adulthood (e.g., ages 15-19).^e.g.,8^ Further, other work has found associations between later-life PLEs and both schizophrenia and mood disorder PGSs.^3^ However, aside from these studies generally examining associations between schizophrenia PGSs and PLEs, no studies, to our knowledge, have examined whether associations are present during childhood and whether PGSs for other phenotypes are associated with PLEs.

Given evidence that PLEs may reflect non-specific indicators of general psychopathology vulnerability that share common genetic risk with a wide range of psychopathologies and related phenotypes (e.g., schizophrenia, depression, ADHD, cognition),^3,9^ genomic vulnerability to broad spectrum psychopathology may confer risk for PLEs in childhood. Further, based on epidemiological studies linking psychosis spectrum symptoms to neurodevelopmental impairments including reduced educational attainment and lower cognition,^10^ polygenic propensity for educational attainment (i.e., which is genetically correlated with cognitive functioning and risk-taking),^11^ may also confer vulnerability for childhood PLEs. Finally, emerging evidence linking PLEs to variation in brain structure, including lower global brain volume,^12^ suggest that brain structure may indirectly link genomic risk to PLE expression.^13^

Using data from genomically-confirmed non-Hispanic children (aged nine-to-ten) of European ancestries who completed the baseline session of the ongoing Adolescent Brain Cognitive Development⍰ (ABCD) study (n=4,650), we examined whether PLEs are associated with PGSs for schizophrenia (SCZ), psychiatric cross-disorder risk (CROSS), PLEs, and educational attainment (EDU). Finally, we probed whether variability in brain structure (i.e., volume, surface area, cortical thickness) and/or proximal PGS behaviors (e.g., cognition for EDU) indirectly associate polygenic scores to PLEs.

## Methods

### Participants

A sample of 11,875 individuals was obtained from the ABCD Study^®^ (Data Release 2.0.1), a large-scale ongoing longitudinal study of children recruited from 22 research sites across the United States (see Supplement for study-wide exclusion criteria).^14^ ABCD Study^®^ data were accessed from the National Institutes of Mental Health Data Archive (see Acknowledgments). Participants who did not pass quality control metrics and those who were not of genomically-confirmed European ancestries were removed leaving a final analytic sample of 4,650 (46.8% female; mean age= 9.93±0.04 years; Supplement).

### Measures

All measures are described in detail within the Supplement.

#### Psychotic-like Experiences

The dimensional total score of the 21-item child-reported Prodromal Questionnaire-Brief Child Version (PQ-BC)^15^ was used to assess PLEs. Further, we generated a binary metric based upon endorsement of at least one significantly distressing PLE (i.e., rating a PLE between three-to-five on a five-point scale, n=972; Table 1).

**Table 1.** Sample Characteristics

| Variable (N) | Significantly Distressing PLEs |  | Total Sample<br>(N=4650) |
| --- | --- | --- | --- |
|  | No Significantly Distressing PLEs<br>(N=3678) | Endorsed Significantly Distressing<br>PLEs<br>(N=972) |  |
| Age in years (4650) | 9.948 ± 7.525 | 9.882 ± 7.351 | 9.934 ± 7.495 |
| Sex, % female (4650) | 1722 (46.8%) | 454 (46.7%) | 2176 (46.8%) |
| Scanner Type (4532) |  |  |  |
| Siemens, % | 2408 (65.5%) | 576 (59.3%) | 2984 (64.2%) |
| Philips, % | 481 (13.1%) | 125 (12.9%) | 606 (13.0%) |
| General Electric, % | 694 (18.9%) | 247 (25.4%) | 942 (20.3%) |
| PLEs (4649) |  |  |  |
| Total PLEs | 1.110 ± 1.875 | 6.180 ± 3.686 | 2.170 ± 3.142 |
| Distressing PLEs | 1.675 ± 2.940 | 16.701 ± 10.036 | 4.816 ± 8.076 |
| Significantly Distressing PLEs | 0±0 | 2.158 ± 1.470 | 0.451 ± 1.105 |
| PGS (4650) |  |  |  |
| SCZ | 3.532x10<sup>-7</sup> ± 1.860x10<sup>-7</sup> | 3.724x10<sup>-7</sup> ± 1.846x10<sup>-7</sup> | 3.572x10<sup>-7</sup> ± 1.855x10<sup>-7</sup> |
| EDU | -1.459x10<sup>-7</sup> ± 9.251x10<sup>-8</sup> | -1.639x10<sup>-7</sup> ± 9.212x10<sup>-8</sup> | -1.497x10<sup>-7</sup> ± 9.271x10<sup>-8</sup> |
| PLE | -1.810x10<sup>-6</sup> ± 6.374x10<sup>-7</sup> | -1.820x10<sup>-6</sup> ± 6.552x10<sup>-7</sup> | -1.808x10<sup>-6</sup> ± 6.411x10<sup>-7</sup> |
| CROSS | 3.029x10<sup>-7</sup> ± 8.535x10<sup>-8</sup> | 3.181x10<sup>-7</sup> ± 8.484x10<sup>-8</sup> | 3.061x10<sup>-7</sup> ± 8.546x10<sup>-8</sup> |
| Global Brain Metrics |  |  |  |
| Intracranial Volume (3914) | 1.560x10<sup>-6</sup> ± 1.303x10<sup>-5</sup> | 1.535x10<sup>-6</sup> ± 1.478x10<sup>-5</sup> | 1.555x10<sup>-6</sup> ± 1.414x10<sup>-5</sup> |
| Total Cortical Volume (4520) | 6.138x10<sup>-5</sup> ± 5.280x10<sup>-4</sup> | 6.066x10<sup>-5</sup> ± 5.448x10<sup>-4</sup> | 6.123x10<sup>-5</sup> ± 5.323x10<sup>-4</sup> |
| Total Subcortical Volume (4520) | 6.145x10<sup>-4</sup> ± 4.755x10<sup>-3</sup> | 6.061x10<sup>-4</sup> ± 4.882x10<sup>-3</sup> | 6.128x10<sup>-4</sup> ± 4.793x10<sup>-3</sup> |
| Total Cortical Thickness (4520) | 2.799 ± 0.106 | 2.785 ± 0.109 | 2.796 ± 0.107 |
| Total Surface Area (4520) | 1.905x10<sup>-5</sup> ± 1.747x10<sup>-4</sup> | 1.892x10<sup>-5</sup> ± 1.761x10<sup>-4</sup> | 1.902x10<sup>-5</sup> ± 1.750x10<sup>-4</sup> |
| Proximal PGS Behaviors |  |  |  |
| Cognitive Scores (4580) | 89.353 ± 7.785 | 86.774 ± 7.968 | 88.814 ± 7.893 |
| General Psychopathology Scores (3120) | -0.012 ± 0.869 | 0.292 ± 0.938 | 0.050 ± 0.892 |

Abbreviations: PLEs=psychotic-like experiences; PGS=polygenic scores; SCZ=Schizophrenia; EDU=Educational Attainment; CROSS=Cross Disorder.

#### Proximal EDU and CROSS PGS Behaviors

Total cognition composite scores assessed using the National Institutes of Health Toolbox Cognitive Battery^16^ and a general psychopathology factor^17^ created using the Child Behavior Checklist,^18^ served as proximal behavioral measures of EDU PGS and CROSS PGS, respectively.

#### Brain Structure

The following global and regional structural MRI metrics were examined: *global:* intracranial, total cortical, and total subcortical volume; total surface area; and total cortical thickness; *regional:* 34 bilateral Desikan cortical regions for surface area, thickness, and volume, as well as 22 Freesurfer segmentation subcortical volume regions (including all 12 bilateral subcortical segmentations).^19^ T1- and T2-weighted structural scans (1mm isotropic) were acquired using 3T scanners (either Siemens, General Electric, or Phillips) with 32-channel head coils.

#### Polygenic Scores

Summary statistics from the most well-powered “discovery” GWASs of SCZ (N=36,989 cases + 113,075 controls),^7^ CROSS (N=232,964 cases [anorexia nervosa, attention-deficit/hyperactivity disorder, autism spectrum disorder, bipolar disorder, major depressive disorder, obsessive-compulsive disorder, schizophrenia, and Tourette’s syndrome] + 494,162 controls),^9^ PLEs (N=127,966),^3^ and EDU (N=766,345)^11^ were used to generate PGS (n=4) within the ABCD Study® dataset. PGS were generated using polygenic risk scores-continuous shrinkage (PRS-CS),^20^ which uses a Bayesian regression framework to include all SNPs in PGS calculation by placing a continuous shrinkage prior on SNP effect sizes; simulation studies show that PRS-CS outperforms other PGS methods.^20^ Analyses using traditional p-value clumping and thresholding produced results consistent with PRS-CS (Supplemental Tables 1-2).

### Statistical Analyses

Individual values on continuous predictor and outcome variables were Winsorized to ±3 SD to minimize the influence of extreme values. All analyses nested data with random intercepts for site (n=22) and family (n=3,874; siblings n=616). The following covariates were included in all analyses: age, sex, genotyping batch, and the first ten ancestrally-informative PCs (described in the Supplement). We used ComBat harmonization (https://github.com/ncullen93/neuroCombat) to estimate and remove scanner model effects from MRI measures. Further, scanner type (i.e., Siemens, Philips, GE) was included as an additional covariate in all brain structure analyses; regional structural analyses further included intracranial volume. Financial adversity and highest parental/caregiver education (described in the Supplement) were included as additional covariates in supplemental analyses. Benjamini-Hochberg False Discovery Rate (FDR) correction was used to adjust for multiple comparisons.

First, we estimated associations between SCZ, PLE, CROSS, and EDU PGS with each PLE metric (i.e., dimensional total score and binary significantly distressing PLEs). FDR correction was used to account for the four PGSs tested.^1^ For CROSS and EDU, *post-hoc* HLMs examined models including measured behaviors most proximal to the PGS, to examine the extent to which available measured outcomes proximal to PGSs accounted for these associations (e.g., the extent to which cognition accounted for the association between EDU PGS and PLE). Second, we estimated associations between reported PLEs and MRI-derived brain structure phenotypes (i.e., total and regional surface area, cortical volume, cortical thickness, and subcortical volume). FDR was used to adjust for multiple testing (i.e., five tests for global MRI metrics, 102 tests for bilateral and average cortical metrics [i.e., Cortical Thickness, Cortical Surface Area, Cortical Volume], and 45 tests for bilateral and average subcortical volume). For any brain structure phenotypes associated with reported PLEs, models subsequently examined the association with each PGS that was associated with reported PLE metrics. Third, we conducted a series of parallel mediation analyses to estimate indirect associations (boot strapped 95% C.I.s) between PGS and reported PLEs through each of the brain structure phenotypes associated with reported PLEs and proximal PGS behaviors entered in parallel. The lme4^21^ (lmer for total PLEs, glmer for binary significantly distress PLEs) and lavaan^22^ R packages were used to conduct hierarchical linear mixed effects regression and nested mediation analyses, respectively.

## Results

### Polygenic Scores and Psychotic-like Experiences

In our sample (n=4,650), 55.33% (n=2,573) of children endorsed at least one reported PLE and 20.90% (n=972) endorsed at least one significantly distressing PLE. *Total PLEs* were associated with higher CROSS and lower EDU PGS (all |βs|>0.045, all ps<0.002, p_FDR_s<0.004; %ΔR^2^s>0.20%), but not SCZ or PLE PGS (all |βs|≤0.015, all ps>0.29; Table 2; Figure 1). When included in the same model, CROSS (all βs>0.042, all ps<0.004) and EDU (all βs>-0.069, all ps=2.96 × 10^−06^) PGS both remained associated with reported PLEs. Endorsement of *Significantly Distressing PLEs* was associated with higher SCZ, PLE, and CROSS PGS as well as lower EDU PGS (all |βs|>0.032, all ps≤0.02, p_FDR_s<0.02; %ΔR^2^s≥0.12%; Table 2; Figure 1). All associations remained similar when further accounting for financial adversity and parental/caregiver education (p_FDR_s<0.03), or when computing PGS using a traditional clustering and thresholding approach (Supplemental Tables 1-2).

**Table 2.** Associations between PLEs and both PGS and Structural Neural Metrics^a^

|  | b [95% CI] | Total PLEs |  |  |  |  | OR [95% CI] | Significant Distress Group |  |  |  |  |
| --- | --- | --- | --- | --- | --- | --- | --- | --- | --- | --- | --- | --- |
| | | $\beta$ | *t* | *p* | *p*<sub>FDR</sub> | %$\Delta R^2$ | | $\beta$ | *Z* | *p* | *p*<sub>FDR</sub> | %$\Delta R^2$ |
| **PGS** |  |  |  |  |  |  |  |  |  |  |  |  |
| Schizophrenia | 2.62x10<sup>5</sup><br>[2.25x10<sup>5</sup>, 7.50x10<sup>5</sup>] | 0.015 | 1.054 | .29 | .36 | 0.022% | 1.110<br>[1.030, 1.200] | **0.038** | **2.736** | **.006** | **.008** | **0.171%** |
| Educational Attainment | **-2.76x10<sup>6</sup>**<br>**[3.74x10<sup>6</sup>, -1.79x10<sup>6</sup>]** | **-0.081** | **-5.561** | **2.85x10<sup>-8</sup>** | **1.14 x10<sup>-7</sup>** | **0.660%** | **0.825**<br>**[0.766, 0.890]** | **-0.072** | **-4.988** | **6.09x10<sup>-7</sup>** | **2.44x10<sup>-6</sup>** | **0.587%** |
| PLEs | 6.21x10<sup>5</sup><br>[6.93x10<sup>5</sup>, 1.94x10<sup>6</sup>] | 0.013 | 0.925 | .36 | .36 | 0.019% | **1.090**<br>**[1.010, 1.170]** | **0.032** | **2.259** | **.02** | **.02** | **0.120%** |
| Cross Disorder | **1.67x10<sup>6</sup>**<br>**[6.18x10<sup>5</sup>, 2.73x10<sup>6</sup>]** | **0.045** | **3.099** | **.002** | **.004** | **0.202%** | **1.190**<br>**[1.110, 1.290]** | **0.066** | **4.600** | **4.23x10<sup>-6</sup>** | **8.46x10<sup>-6</sup>** | **0.505%** |
| **Neural Metrics** |  |  |  |  |  |  |  |  |  |  |  |  |
| **Global Metrics** |  |  |  |  |  |  |  |  |  |  |  |  |
| Intracranial Volume | **-1.48x10<sup>-6</sup>**<br>**[-2.36x10<sup>-6</sup>, -5.93x10<sup>-7</sup>]** | **-0.065** | **-3.274** | **.001** | **.001** | **0.062%** | **0.858**<br>**[0.774, 0.950]** | **-0.055** | **-2.935** | **.003** | **.004** | **0.372%** |
| Total Cortical Volume | **-4.03 x10<sup>-6</sup>**<br>**[-5.96x10<sup>-6</sup>, -2.10x10<sup>-6</sup>]** | **-0.067** | **-4.091** | **4.37x10<sup>-5</sup>** | **2.19x10<sup>-4</sup>** | **0.380%** | **0.847**<br>**[0.779, 0.921]** | **-0.061** | **-3.865** | **1.11x10<sup>-4</sup>** | **2.78x10<sup>-4</sup>** | **0.332%** |
| Total Subcortical Volume | **-3.83x10<sup>-5</sup>**<br>**[-6.00x10<sup>-5</sup>, -1.67x10<sup>-5</sup>]** | **-0.057** | **-3.466** | **5.33x10<sup>-4</sup>** | **1.25x10<sup>-3</sup>** | **0.271%** | **0.825**<br>**[0.757, 0.898]** | **-0.070** | **-4.201** | **9.78x10<sup>-6</sup>** | **4.89x10<sup>-5</sup>** | **0.442%** |
| Total Cortical Thickness | **-1.21 [-2.192, -0.235]** | **-0.041** | **-2.430** | **.02** | **.02** | **0.161%** | **0.915**<br>**[0.841, 0.997]** | **-0.032** | **-1.868** | **.04** | **.04** | **0.069%** |
| Total Surface Area | **-1.08x10<sup>-5</sup>**<br>**[-1.71x10<sup>-5</sup>, -4.54x10<sup>-6</sup>]** | **-0.058** | **-3.374** | **7.47x10<sup>-4</sup>** | **1.25x10<sup>-3</sup>** | **0.247%** | **0.858**<br>**[0.785, 0.938]** | **-0.057** | **-3.231** | **7.74x10<sup>-4</sup>** | **.001** | **0.241%** |
| **Regional Metrics** |  |  |  |  |  |  |  |  |  |  |  |  |
| Average Inferior Parietal Thickness | **-1.47 [-2.301, -0.646]** | **-0.069** | **-3.490** | **4.89x10<sup>-4</sup>** | **.03** | **0.210%** | 0.886<br>[0.7950, 0.998] | -0.043 | -2.185 | .03 | .27 | 0.122% |
| Right Inferior Parietal Thickness | **-1.35 [-2.117, -0.585]** | **-0.067** | **-3.457** | **5.53x10<sup>-4</sup>** | **.03** | **0.233%** | 0.888<br>[0.803, 0.982] | -0.045 | -2.324 | .02 | .27 | 0.138% |

Abbreviations: PLEs=psychotic-like experiences; PGS=polygenic scores; b=unstandardized beta coefficient; 95% CI=95% confidence interval; $\beta$ =standardized beta coefficient; *t*=t-test test statistic; *p*=*p*-value; *p*<sub>FDR</sub>=False Discovery Rate-corrected *p*; % $\Delta R^2$ =percentage change in marginal proportion of variance explained, calculated as the difference in $R^2$ between the current model versus a model excluding the predictor of interest, converted to a percentage; OR=odds ratio; *Z*=*Z* statistic.
<sup>a</sup>*p*<sub>FDR</sub><.05 models are in bold. All PGS (i.e., SCZ, educational attainment, cross disorder, PLE) results remained consistent with accounting for financial adversity and parental education.

**Figure 1.**
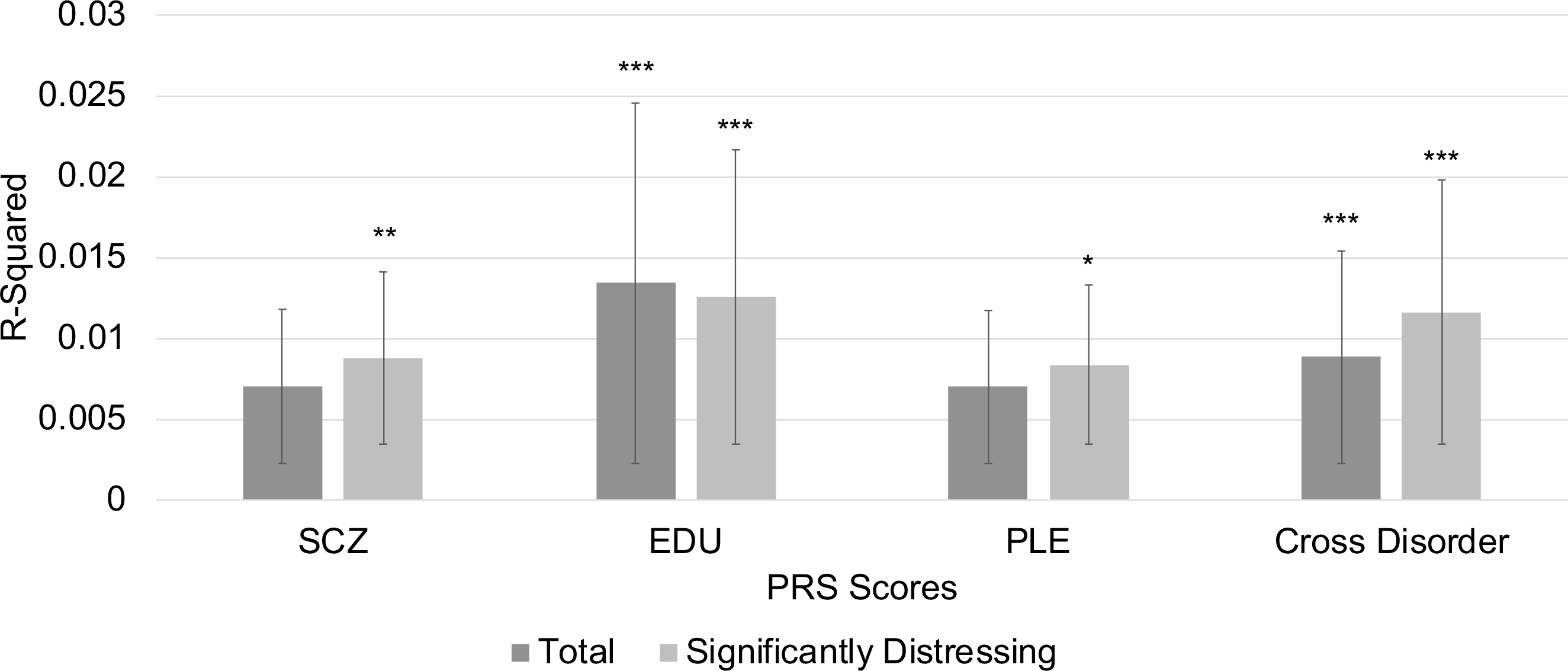
Proportion of variance explained (R-squared) by each of the different PGS scores (SCZ=Schizophrenia; EDU =Educational Attainment; PLE=Psychotic-like experiences; CROSS=Cross Disorder) for both Total and Significantly Distressing PLEs. Error bars represent the 95% confidence interval, * p_FDR_<0.05, ** p_FDR_<0.01, *** p_FDR_<0.001.

#### Consideration of Proximal EDU and CROSS PGS Behaviors: Cognition and General Psychopathology

Measured cognition was negatively associated with *Total PLEs* and endorsement of *Significantly Distressing PLEs* and positively associated with EDU PGS (|βs|≥0.131, all ps<2.00 × 10^−16^, %ΔR^2^s≥1.55%; Supplemental Table 3; Supplemental Figures 1-2). Cognitive performance accounted for 32.3-32.8% of the association between EDU PGS and PLE metrics (Supplemental Table 4), though associations between EDU PGS and reported PLE metrics remained, even when considering cognition (βs>-0.047, ps<0.002; %ΔR^2^s≥0.58%; Supplemental Table 5).

**Table 3.** Associations between PGS and Structural Neural Metrics^a^

|  | Educational Attainment PGS |  |  |  |  |  | Cross Disorder PGS |  |  |  |  |  |
| --- | --- | --- | --- | --- | --- | --- | --- | --- | --- | --- | --- | --- |
| | b [95% CI] | $\beta$ | $t$ | $p$ | $p_{FDR}$ | %$\Delta R^2$ | b [95% CI] | $\beta$ | $t$ | $p$ | $p_{FDR}$ | %$\Delta R^2$ |
| **Global Structural Metrics** |  |  |  |  |  |  |  |  |  |  |  |  |
| Intracranial Volume | 1.08x10<sup>11</sup><br>[7.06x10<sup>10</sup>, 1.46x10<sup>11</sup>] | **0.073** | **5.640** | **1.82x10<sup>-8</sup>** | **3.03x10<sup>-8</sup>** | **0.705%** | 1.98x10<sup>10</sup><br>[-2.09x10<sup>10</sup>, 6.05x10<sup>10</sup>] | 0.012 | 0.953 | .34 | .43 | 0.005% |
| Total Cortical Volume | 4.93x10<sup>10</sup><br>[3.43x10<sup>10</sup>, 6.43x10<sup>10</sup>] | **0.088** | **6.440** | **1.32x10<sup>-10</sup>** | **6.60x10<sup>-10</sup>** | **0.825%** | 1.43x10<sup>10</sup><br>[-1.97x10<sup>9</sup>, 3.06x10<sup>10</sup>] | 0.023 | 1.723 | .09 | .21 | 0.040% |
| Total Subcortical Volume | 2.91x10<sup>9</sup><br>[1.57x10<sup>9</sup>, 4.24x10<sup>9</sup>] | **0.058** | **4.257** | **2.12x10<sup>-5</sup>** | **2.65x10<sup>-5</sup>** | **0.353%** | 4.83x10<sup>8</sup><br>[-9.67x10<sup>8</sup>, 1.93x10<sup>9</sup>] | 0.009 | 0.653 | .51 | .51 | 0.000% |
| Total Cortical Thickness | 1.59x10<sup>4</sup><br>[-1.39x10<sup>4</sup>, 4.58 x10<sup>4</sup>] | 0.014 | 1.045 | .30 | .30 | 0.066% | -2.35x10<sup>4</sup><br>[-5.58x10<sup>4</sup>, 8.80x10<sup>4</sup>] | -0.019 | -1.426 | .15 | .26 | 0.025% |
| Total Surface Area | 1.44x10<sup>10</sup><br>[9.82x10<sup>9</sup>, 1.90x10<sup>10</sup>] | **0.079** | **6.139** | **9.04x10<sup>-10</sup>** | **2.26x10<sup>-9</sup>** | **0.568%** | **7.25x10<sup>9</sup>**<br>**[2.26x10<sup>9</sup>, 1.22x10<sup>10</sup>]** | **0.037** | **2.849** | **.004** | **.02** | **0.110%** |
| **Regional Metrics** |  |  |  |  |  |  |  |  |  |  |  |  |
| Average Inferior Parietal Thickness | 6.71x10<sup>3</sup><br>[-3.15x10<sup>4</sup>, 4.49x10<sup>4</sup>] | 0.004 | 0.344 | .73 | .73 | 0.026% | -1.16x10<sup>4</sup> [-5.30x10<sup>4</sup>, 2.98x10<sup>4</sup>] | -0.007 | -0.548 | .58 | .58 | 0.005% |
| Right Inferior Parietal Thickness | -1.14x10<sup>4</sup><br>[-5.24x10<sup>4</sup>, 2.95x10<sup>4</sup>] | -0.007 | -0.547 | .58 | .73 | 0.024% | -2.18x10<sup>4</sup><br>[-6.61x10<sup>4</sup>, 2.25x10<sup>4</sup>] | -0.012 | -0.964 | .34 | .58 | 0.003% |

Abbreviations: PLEs=psychotic-like experiences; PGS=polygenic scores; b=unstandardized beta coefficient; 95% CI=95% confidence interval; $\beta$ =standardized beta coefficient; $t$ =t-test test statistic; $p$ =p-value; $p_{FDR}$ =False Discovery Rate-corrected $p$ ; % $\Delta R^2$ =percentage change in marginal proportion of variance explained, calculated as the difference in $R^2$ between the current model versus a model excluding the predictor of interest, converted to a percentage; OR=odds ratio; Z=Z statistic.
<sup>a</sup> $p_{FDR} < .05$ models are in bold.

The general psychopathology factor was positively associated with *Total PLEs* and endorsement of *Significantly Distressing PLEs* as well as CROSS PGS (all βs≥0.057, all ps≤0.001, %ΔR^2^s>0.29%; Supplemental Table 3; Supplemental Figures 1-2). General psychopathology accounted for between 11.3-17.8% of the association between CROSS PGS and reported PLEs, though these remained associated with each other (*Total*: β=0.034, p=0.048, ΔR^2^=0.24%; *Significantly Distressing PLEs*: β=0.056, p=0.002, %ΔR^2^=0.72%; Supplemental Table 5).

### Psychotic-like Experiences and Brain Structure

Greater reported PLEs (both *Total* and endorsement of *Significantly Distressing PLEs*) were associated with lower intracranial volume, total cortical volume, total subcortical volume, total cortical thickness, and total surface area (all βs<-0.032, all ps<0.04, all p_FDR_s<0.04; all %ΔR^2^s>0.06; Table 2; Figure 2). When examining individual structural MRI regions for volume, surface area, and thickness, greater *Total PLEs* were associated with lower average and right inferior parietal cortical thickness (both βs>-0.067, both ps<5.53×10^−4^, both p_FDR_s=0.03; both %ΔR^2^s>0.21%; Table 2; Figure 2). No other individual structural MRI regions passed FDR correction (see Supplemental Tables 6-13).

**Figure 2.**
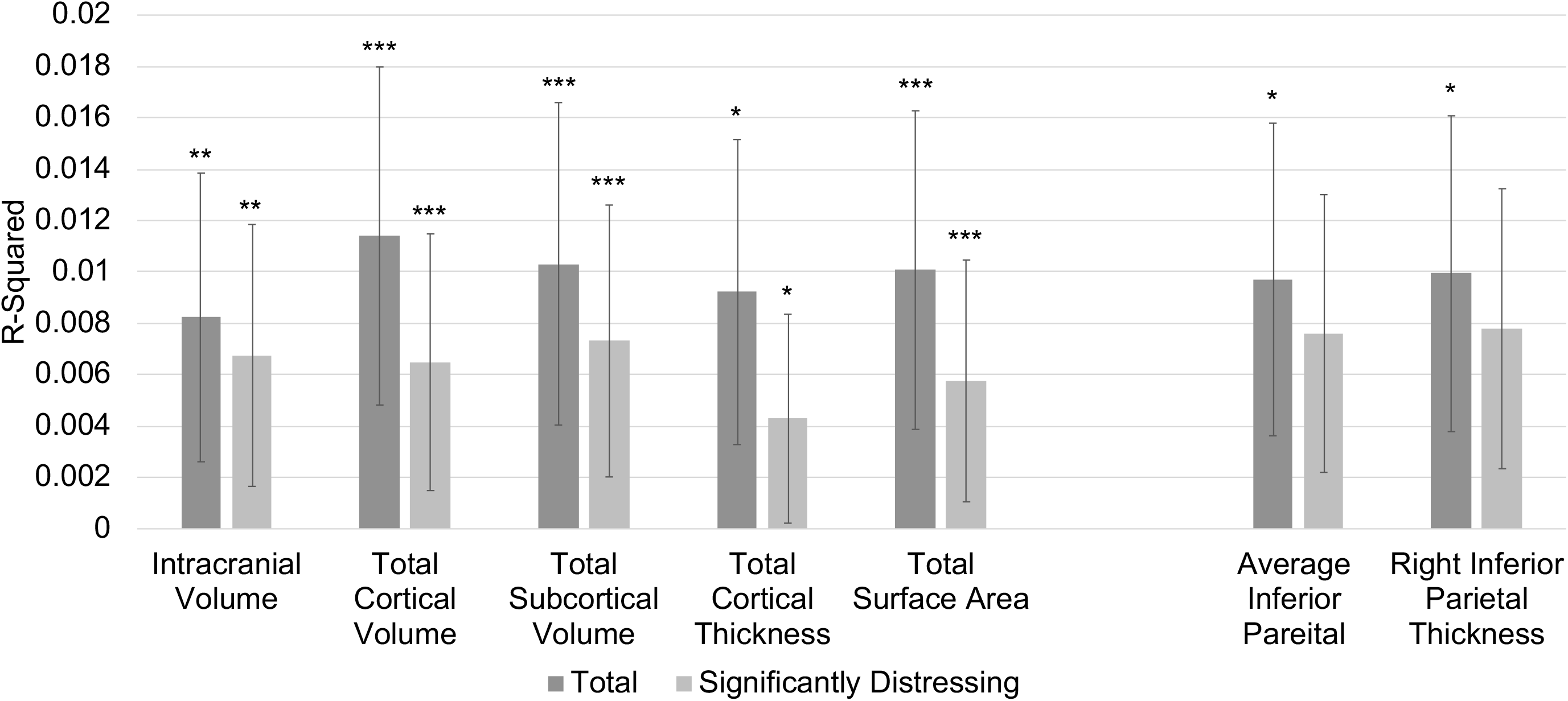
Proportion of variance explained (R-squared) by each of the significant global (intracranial volume, total cortical volume, total subcortical volume, surface area) and regional (inferior parietal thickness) MRI metrics for both Total and Significantly Distressing PLEs. Error bars represent the 95% confidence interval, * p_FDR_<0.05, ** p_FDR_<0.01, *** p_FDR_<0.001.

### Mediation Analyses

#### Educational Attainment Polygenic Scores

EDU PGS were associated with brain structure phenotypes associated with reported PLEs (i.e., intracranial volume, total cortical volume, total subcortical volume, total cortical surface area; all βs>0.058, all ps<2.65E-05; Table 3), except global and inferior parietal cortical thickness (all |βs|≤0.014, all ps>0.30). A series of individual mediational models (n=4) examined whether each brain structure phenotype associated with both EDU PGS and PLEs (i.e., intracranial volume, total cortical volume, total subcortical volume, total cortical surface area) indirectly linked EDU PGS to *Total PLEs* alongside cognitive performance in parallel. All volume metrics (i.e., intracranial volume, total cortical volume, and total subcortical volume) partially mediated the association between EDU PGS and *Total PLEs* (all indirect effect [path *a*b*] bias-corrected 95% confidence intervals [CI] within - 0.012 to −0.001; proportion mediated:3.33%-8.79%; Figure 3a-c). There was not a strong indirect association between PGS and *Total PLEs* through total surface area (indirect effect 95% CI: −0.004 to 0.002; Figure 3d). Cognition uniquely partially mediated the association between EDU PGS and *Total PLEs* in each model (all within 95% C.I.: [CI], −0.038 to −0.020; proportion mediated:28.57-32.22%; Figure 3). For all models, similar results were found when endorsement of *Significantly Distressing PLEs* was the outcome instead of *Total PLEs* (Supplement). A single parallel mediation model with all neural metrics entered as simultaneous parallel mediators is reported in the Supplement.

**Figure 3.**
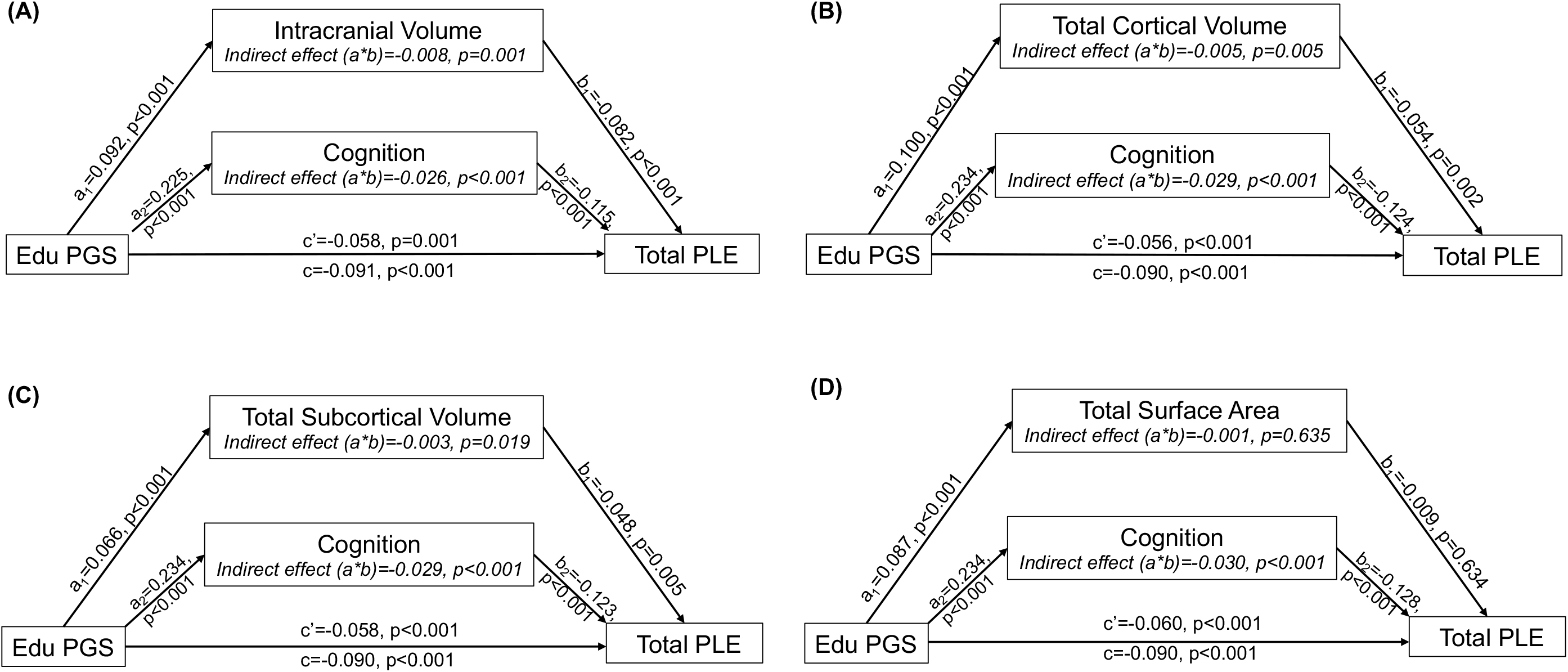
Depiction of a series of parallel mediation models, examining evidence for each neural metric ((A) intracranial, (B) cortical volume, (C) subcortical volume, and (D) surface area) indirectly linking Educational Attainment PGS (EDU PGS) and total psychotic-like experiences (Total PLEs) alongside cognitive performance in parallel. Covariates (i.e., age, sex, genotyping batch, and the first ten ancestrally-informative PCs) were included in all models. Each parallel mediation model depiction includes unstandardized regression coefficients, showing the association between EDU PGS, each neural metric and cognition, and Total PLEs.

#### Schizophrenia and Cross Disorder Polygenic Scores

SCZ and PLE PGS were not associated with reported PLE-related global or regional brain structure measures (Supplemental Table 14). Although total surface area was associated with CROSS PGS (β=0.037, p=0.004, p_FDR_=0.02; %ΔR^2^=0.11; Table 3), it did not mediate associations between CROSS PGS and reported PLEs (95% CI: −0.005 to 0.000; Supplement). No other reported PLE-related brain phenotypes were associated with CROSS PGS (Table 3).

## Discussion

Here, we show that psychotic-like experiences (PLEs) in middle childhood (n=4,650) are associated with GWAS-derived indices of polygenic risk (all %ΔR^2^s=0.120-0.660%) as well as potential putative intermediary neural and behavioral phenotypes that may partially underlie these associations (all %ΔR^2^s=0.062-2.85%). Genomic liability for lower educational attainment and higher psychopathology (i.e., general psychopathology, schizophrenia, late-life PLEs) were associated with more PLEs among children (Figure 1). There was divergence in the strength of associations across phenotypic definitions of PLEs (i.e., broad–total PLEs, versus severe–presence of significantly distressing PLEs). Specifically, genomic liability for broad spectrum psychopathology (CROSS PGS) and educational attainment (EDU PGS) were associated with both PLEs measures; SCZ and late-life PLE PGS were associated only with the presence of significantly distressing PLEs. Broadly, this divergence mirrors phenotypic observations that PLEs may portend broad psychopathology vulnerability,^1^ while severe expressions of PLEs may foreshadow psychosis risk.^23^ However, the direction of association and estimated effect size, despite differences in significance, were similar across phenotypic PLE measures (i.e., broad and severe; Figure 1). Finally, reported PLEs were associated with lower global (e.g., intracranial volume) and regional (i.e., inferior parietal thickness was associated with broad PLEs) brain structure metrics (Figure 2), with evidence that lower volume (i.e., ICV, total cortical and total subcortical) indirectly links EDU PGS to reported PLEs alongside cognition (Figure 3). Collectively, these results show that polygenic scores derived from GWAS of adult samples can generalize to indices of risk among children. Because PLEs can develop in children with a negative family history of psychotic disorders, they may, eventually, prove useful indices of risk for psychopathology beyond familial history. As larger GWASs of both precise and broad phenotypes continue to emerge, polygenic risk indices may help target individuals for preventative therapies (e.g., cognitive behavioral interventions) and further help disarticulate the biological and behavioral mechanisms underlying psychiatric risk that may ultimately contribute to advances in treatment and nosology.

### Polygenic Propensity for Education Attainment

Polygenic scores for lower educational attainment were the most robust PGS predictor of both total PLEs and significantly distressing PLEs. These findings suggest that prior reports linking EDU PGS to severe psychosis among clinical patients^24^ may generalize to earlier markers of psychosis spectrum symptoms during middle childhood. That cognition accounted for a large portion (28.57-32.22% of variance) of the association between educational attainment and reported PLEs aligns with evidence that premorbid cognition prospectively predicts PLEs and psychopathology, including schizophrenia,^10^ and suggests that such vulnerability may be partially genomic in origin. However, other work has failed to find a strong genetic correlation between later-life PLEs and intelligence;^3^ it is possible that genomic associations between PLEs and cognition may differ across the life course and be more correlated during childhood with divergence in later life (e.g., PLEs related to cognitive decline and/or dementia^25^).

Supportive of neurodevelopmental models of psychosis spectrum disorders which posit that brain differences underlie cognition-related vulnerability for schizophrenia, we found evidence that brain volume accounts for a portion of the association between educational attainment PGS and reported PLEs (3.22-8.79%; Supplemental Table 4; Figure 3). However, given that these neural metrics are less proximal to educational attainment polygenic scores than cognition, it is unsurprising that associations for cognition were larger than associations for brain structure.^26^ Notably, these associations between reported PLEs and global volume reductions are consistent with previous research examining volumetric alterations.^12^ That lower global volume may indirectly link educational attainment PGS and reported PLEs is consistent with the notion that genomic liability for lower cognitive functioning may be associated with altered neural maturational processes, which may contribute to the development or maintenance of psychosis spectrum symptoms.^27^

### General Psychopathology, Schizophrenia, and PLE Polygenic Risk

Genomic liability for broad spectrum psychopathology (i.e., CROSS PGS) was associated with broadly defined as well as severe PLEs, while polygenic scores for SCZ and PLEs were only associated with severe PLEs. These findings add to developing evidence from phenotypic research that PLEs represent transdiagnostic harbingers of broad-spectrum psychopathology as opposed to psychosis-specific risk, while more severe forms of PLEs that elicit distress may be more uniquely associated with psychosis-specific liability.^1,2^ This is consistent with prior research finding stronger associations between SCZ PGS and distressing PLEs; however, this previous work also found associations between later-life broadly defined PLEs and SCZ PGS.^3^ These findings also align with evidence that polygenic and phenotypic psychopathology associations may be more non-specific in middle childhood and potentially become increasingly specific with increased severity (e.g., with more clinically significant PLEs) and/or maturation (e.g., in adolescence).^28^ We did not find strong evidence of associations between broadly defined PLEs with polygenic risk for later life PLEs, although there were associations between later life PLEs PGS and more severe PLEs. It is entirely possible that PLEs during *middle/late life* (the basis of the PLE PGS score used here) are somewhat etiologically distinct from broadly defined *childhood* PLEs. Further, it is important to consider that the CROSS disorder PGS was generated using GWASs of eight psychopathologies (i.e., schizophrenia, bipolar disorder, major depressive disorder, attention-deficit/hyperactivity disorder, autism spectrum disorder, obsessive-compulsive disorder, anorexia nervosa, Tourette syndrome), of which psychosis spectrum disorders (i.e., schizophrenia and bipolar disorder) were the most well-powered. Thus, CROSS PGS effects are potentially driven by schizophrenia and its pleiotropy. Alongside evidence of similar effect size estimates across both PLE measures within PGS, it is plausible that with better powered discovery GWAS and target samples, PLEs defined broadly and with regard to distress will show similar relationships.

## Limitations

It is important to consider major limitations of this study while interpreting these findings. First, the generalizability of these findings are limited, as we restricted analyses to those of genomically-confirmed European ancestries due to the sample compositions of the discovery GWASs and evidence that polygenic risk does not translate across ancestries.^29^ Second, reported PLEs were assessed only via self-report and were not corroborated with clinical interview. However, initial evidence from the ABCD Study® suggests that PLEs can be validly assessed using the PQ-BC in a middle childhood sample, as evidenced by associations with family history of psychosis, impaired cognition, and other indices of psychopathology.^15^ Third, and consistent with expectations and prior PGS studies, the effects reported are generally small (for associations with reported PLEs, |βs|<.09) and suggest that PGS in their current form have little direct applied clinical utility for childhood PLEs. As “discovery” GWASs continue to grow, it will be critical to acquire additional GWASs datasets across development, including in childhood, to examine genetic correlations for the same phenotype across ages as well as differential associations with psychopathology and structural neural metrics. Fourth, our mediational analyses are non-experimental and based on cross-sectional data. While these analyses provide empirical evidence consistent with putative gene-brain-behavior mechanisms underlying childhood PLE risk, they should not be interpreted by themselves to imply causation. For example, it remains plausible that the presence of PLEs could influence brain structure. Lastly, it is important to note that the GWASs have differential power to detect effects (e.g., the EDU GWAS was based on 766,345 individuals, versus the PLEs GWAS was based on 127,966 individuals). These differences in power present challenges for interpreting across different PGSs.

## Conclusions

Broadly, GWAS-based polygenic scores are associated with indices of psychopathology and PLEs during middle childhood. Polygenic liability to educational attainment was the most robust predictor of both broadly defined PLEs as well as severe PLEs, and there was evidence that these associations may be partially mediated by cognitive performance and brain structure. Polygenic score associations mirrored phenotypic evidence that broadly defined PLEs are associated with severe broad-spectrum psychopathology risk while more severe PLEs may index psychosis liability. Taken together, the current study documents that GWAS-derived polygenic scores index psychopathology and psychosis vulnerability in children. The PGSs do not have current clinical utility but support the identification of putative intermediate biological and behavioral mechanisms through which genomic risk for psychopathology emerges.

## Contributors

NRK, RB, and DMB developed the research questions. NRK, SEP, ECJ, and ASH conducted analyses. RB, DMB, AA, WKT, and DAAB provided critical revision of the manuscript for important intellectual content. NRK, RB, SEP, and ECJ had full access to all data and take responsibility for the integrity of the data and accuracy of analyses. All authors have approved the final version of the manuscript.

## Data Availability

Data used in the preparation of this article were obtained from the Adolescent Brain Cognitive Development (ABCD) Study (https://abcdstudy.org), held in the NIMH Data Archive (NDA).

## Acknowledgments

Data used in the preparation of this article were obtained from the Adolescent Brain Cognitive Development (ABCD) Study (https://abcdstudy.org), held in the NIMH Data Archive (NDA). This is a multisite, longitudinal study designed to recruit more than 11,500 children age 9-10 and follow them over 10 years into early adulthood. The ABCD Study is supported by the National Institutes of Health and additional federal partners under award numbers U01DA041022, U01DA041025, U01DA041028, U01DA041048, U01DA041089, U01DA041093, U01DA041106, U01DA041117, U01DA041120, U01DA041134, U01DA041148, U01DA041156, U01DA041174, U24DA041123, and U24DA041147. A full list of supporters is available at https://abcdstudy.org/nih-collaborators. A listing of participating sites and a complete listing of the study investigators can be found at https://abcdstudy.org/principal-investigators.html. ABCD consortium investigators designed and implemented the study and/or provided data but did not necessarily participate in analysis or writing of this report. This manuscript reflects the views of the authors and may not reflect the opinions or views of the NIH or ABCD consortium investigators.

The ABCD data repository grows and changes over time. The ABCD data used in this report came from DOI 10.15154/1460410.

The authors thank Drs. Michelini and Kotov for the creation of the general psychopathology scores.

This work was supported by National Institute of Health grants U01 DA041120 (DMB), MH014677 (NRK), MH109532, DA032573 (AA), F32 AA027435 (ECJ), T32-DA007261 (ASH), R01-AG045231, R01-HD083614, R01-AG052564, R21-AA027827, R01-DA046224 (RB).

## Footnotes

1 Given the strong correlation (r=.66) between PLE total score and PLE significant distress group, we did not adjust for multiple testing for these 2 phenotypes.

